# Angiotensin 1-7 in severe COVID-19 patients: a phase 1 clinical trial

**DOI:** 10.1101/2022.09.15.22279897

**Authors:** Ana Luiza Valle Martins, Filippo Annoni, Filipe Alex da Silva, Lucas Bolais-Ramos, Gisele Capanema de Oliveira, Alana Helen dos Santos Matos, Maria Cecília Jardim Heyden, Beatriz Dias Pinheiro, Natália Abdo Rodrigues, Danilo Augusto Alves Pereira, Mirella Monique Lana Diniz, Thuanny Granato Fonseca Silva, Alexandre Carvalho Cardoso, Juliana Carvalho Martins, Daisy Motta-Santos, Maria José Campagnole-Santos, Thiago Verano-Braga, Fabio Silvio Taccone, Robson Augusto Souza dos Santos

## Abstract

**Background:** The coronavirus-related disease (COVID-19) is mainly characterized by a respiratory involvement, with few available therapeutics for critically cases. The renin-angiotensin system (RAS) has a relevant role in the pathogenesis of COVID-19, as the virus enter host’s cells via the angiotensin-converting enzyme 2 (ACE2) and RAS disequilibrium promote inflammation and fibrosis. Exogenous angiotensin-(1-7) might modulate RAS in COVID-19 patients; however, no data on its safety are available in this setting.

**Methods:** This investigator-initiated, open label, phase I clinical trial was conducted to test the safety of intravenous administration of Angiotensin-(1-7) in severe COVID-19 patients admitted in two intensive care units (ICU) in Belo Horizonte, Brazil. In addition to standard of care, intravenous administration of Angiotensin-(1-7) was started at 5 mcg/Kg*day and increased to 10 mcg/Kg*day after 24 hours and continued for a maximum of 7 days or until ICU discharge. The rate of serious adverse events (SAEs) served as the primary outcome of the study.

**Results:** Between August and December 2020, 28 patients were included (mean age of 55.8±12.0 years). All but one patient underwent dose escalation after 24 hours and 8 (28.5%) received the treatment until day 7. No significant differences in mean blood pressure and heart rate were observed before and after the initiation of the drug. During the period of intervention, 5/28 (17.8%) patients required vasopressors, 4 at low dose norepinephrine (i.e. <0.05 mcg/kg*min), while one patient required higher doses because of septic shock. One patient presented with sinus bradycardia, which was considered possibly related to the study drug and resolved after discontinuation. Six patients (21.4%) died before ICU discharge.

**Conclusions:** Intravenous infusion of Angiotensin-(1-7) up to 10 mcg/Kg*day was safe in severe COVID-19 patients and could represent a potential therapeutic strategy in this setting.

**Trial Registration:** Registro Brasileiro de Ensaios Clínicos, UTN code: U1111-1255-7167, registered on 08/05/2020; ClinicalTrials.gov Identifier: NCT04633772; retrospectively registered on November 18 2020

## Introduction

The current pandemic induced by the novel Coronavirus (SARS-CoV-2) [1] has been responsible for a high death toll worldwide, reaching more than 6 million by May 2022 [3]. The Coronavirus Disease (COVID-19) can manifest with a large spectrum or respiratory involvement, ranging from mild and self-limiting disease to fast progressive bilateral pneumonia, eventually leading to death [4-7]. Up to date, only few therapeutic agents (i.e., corticosteroids, anti-IL6 agents) have been recommended in severe COVID-19 patients hospitalized in intensive care units (ICUs), all of them being immuno-modulatory and anti-inflammatory drugs [8,9]

The disease pathophysiology remains however complex, and the development of new and affordable medical treatments is mandatory to limit the burden of COVID-19 and increase the possibilities to treat more severe patients. In particular, the role of the renin angiotensin system (RAS) and its relationship with coronaviruses infections has been highlighted since years. The Angiotensin-converting enzyme 2 (ACE2) proteins serves as the cellular binding site for the spike proteins of SARS-CoV-2, which leads to the virus entrance in the cells and viral replication [10].

Angiotensinogen, a protein primarily synthesized in the liver, is transformed into Angiotensin I (Ang I) by renin and is then cleaved into Angiotensin II (Ang II) by the dipeptidyl carboxypeptidase, angiotensin converting enzyme (ACE). Ang II is then capable of triggering different responses in multiple tissues by binding to its specific receptors, named AT1 and AT2. Both Ang I and Ang II could be processed by endopeptidases and ACE2 to Angiotensin-(1-7) (Ang-(1-7)), which binds to a specific receptor (Mas, MasR) to trigger a series of biological responses (Figure 1) [11, 12]. Interestingly, the ACE2/Ang-(1-7)/MasR pathway appears to counterbalance the effects of the ACE/Ang II/AT1, leading to a more precise modulation of several biological processes, such as inflammatory response, tissue fibrosis, blood pressure regulation, renal function, angiogenesis, endocrine and hormonal functions [11,12].

**Figure 1:**
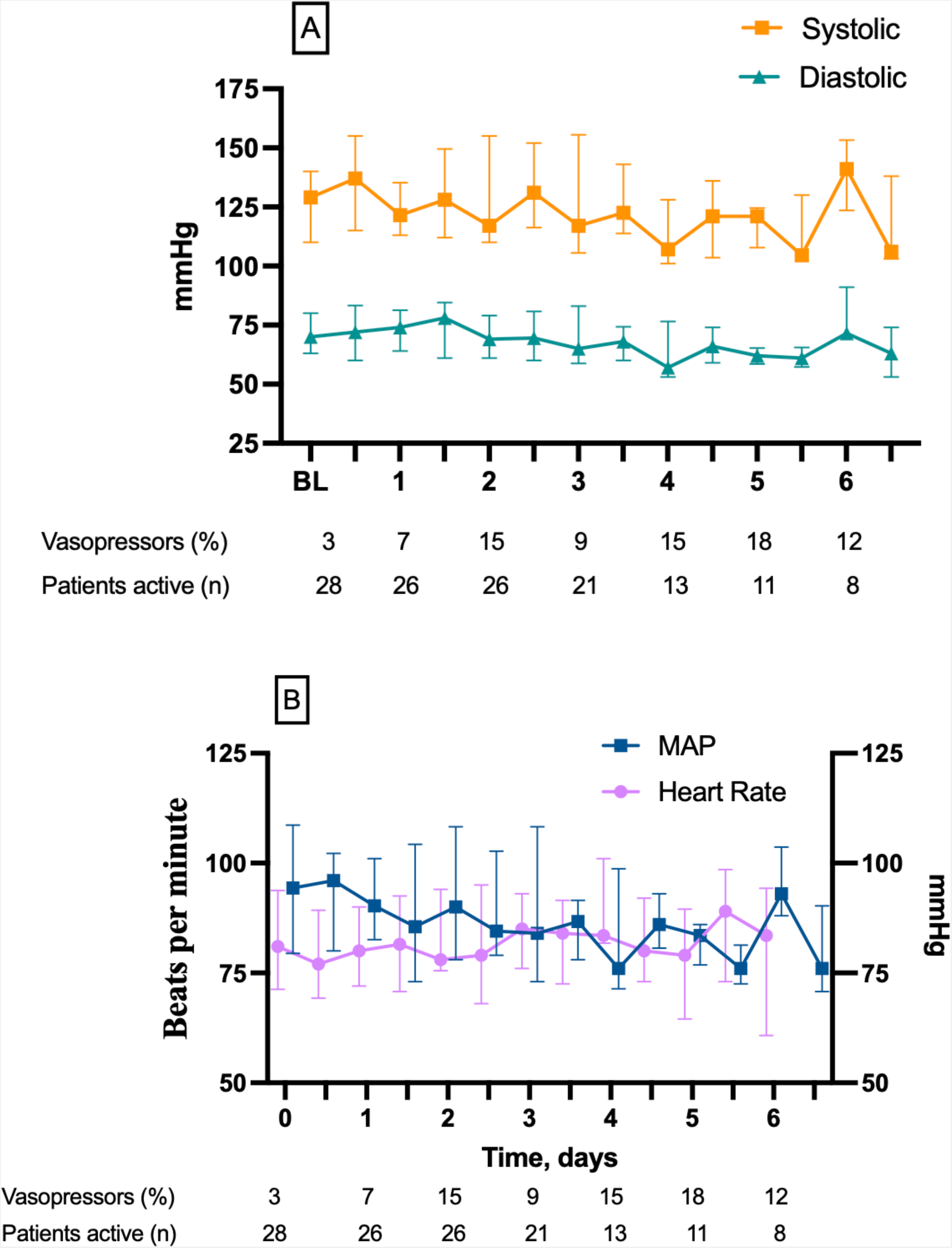
Hemodynamic parameters: systolic and diastolic arterial blood pressure (A) and hearth rate and mean arterial pressure (B). Symbols represents medians and bars represents interquartile range.

During COVID-19 course, ACE2 becomes severely dysregulated, provoking a profound imbalance of the RAS [13]. A large amount of pre-clinical and experimental evidence indicates that the activation of the lung-based RAS is involved in the pathophysiology of pulmonary inflammation [12-17], while the ACE2/Ang-(1-7)/Mas receptor pathway exhibits anti-inflammatory effects in different pulmonary diseases [12-20]. Moreover, Ang-(1-7) mediated effects extend beyond the respiratory and cardiovascular system, i.e., antithrombotic effects [19], reduced muscle atrophy [20] and oxidative stress [12, 23]. In COVID-19 patients, plasmatic Ang-(1-7) levels were slightly increased, and Ang II reduced compared to healthy subjects [24,25]; however, lung RAS dysregulation as well as a relative Ang-(1-7) insufficiency have not been specifically assessed. Although Ang-(1-7) has been suggested as a possible therapeutic agent in COVID-19 [24,25], no study has evaluated the safety of intravenous administration of Ang-(1-7) in this setting.

The aim of this phase I study was therefore to assess the safety of a dose-escalating intravenous administration of Ang-(1-7) in severe COVID-19 patients admitted into the ICU.

## Methods

### Study design and population

This phase I, investigator-initiated, open-label clinical trial was conducted in two ICUs (Mater Dei Hospital and Eduardo de Menezes Hospital) in Belo Horizonte, Brazil. Eligible patients were adult (>18 and <80 years of age) patients with confirmed SARS-CoV-2 infection requiring an ICU admission for respiratory impairment. Patients with a diagnosis of cancer, requiring moderate to high-dose vasopressors (i.e. norepinephrine >0.5 mcg/kg/min), immunocompromised, with active limitations of care, with cardiac failure as the main cause of respiratory failure, suffering from idiopathic lung fibrosis, chronic dialysis, decompensated liver cirrhosis, treated with oxygen therapy at home, pregnant women or if included in any other interventional trial, were excluded. The study protocol was approved by the Ethics Committees of participating centers. Written informed consent was obtained from a legal representative or the patient, depending on the circumstances, before inclusion. All recorded data were collected and managed using REDCap electronic data capture tool. No commercial funding was obtained for this study.

### Study Intervention

Included patients received, in addition to standard treatment according to guidelines of the Brazilian Ministry of Health for patients with Coronavirus infection [37], a continuous intravenous administration of Ang-(1-7) (GMP grade peptide donated by BCN Peptides, Barcelona, Spain and formulated for intravenous infusion by Citopharma Manipulação de Medicamentos Especiais LTDA - Belo Horizonte, Minas Gerais, Brazil) diluted in saline solution 0.9% at the initial dose of 5 mcg/Kg*day, which was increased after 24 hours to 10 mcg/Kg*day for a maximum duration of 7 days or until clinical improvement with ICU discharge or death, whichever came first. Other interventions, including use of mechanical ventilation, renal replacement therapy, vasopressors, or extra-corporeal membrane oxygenation were decided by the attending physician. All decisions about withdrawal of life-sustaining therapies were also at the discretion of the treating physician, and independent from the study protocol.

### Study Outcomes

The primary outcome was the occurrence of Severe Adverse Events (SAEs), recorded after the initiation of therapy and during the time of drug administration; their occurrence was immediately notified to the Data Safety Monitoring Committee (DSMC) for evaluation. SAEs were defined according to the FDA definition [38] and recorded as related or not with the study medication. Since the peptide could decrease vascular tone, we anticipated that the main expected SAE could have been the occurrence of hypotension.

Secondary outcome included: a) ICU length of stay; b) ICU mortality; a) the number of Oxygen Free Days (OFDs) by day 28, defined as the number of days the patient was free from any oxygen supplementation after the inclusion until day 28; b) ICU free days; c) hospital length of stay; d) ventilator free days by day 28 (i.e. the number of days the patients was liberated from mechanical ventilation after inclusion until day 28; e) secondary infections; f) needs for vasopressors; g) incidence of clinically relevant deep vein thrombosis; h) Ang I, Ang II, Ang-(1-5) and Ang-(1-7) plasmatic concentrations, at baseline (T0) and 3 hours (T1), 24 hours (T2) and 72 hours (T3) after the initiation of infusion. A mass spectrometry-based approach was used to quantify these RAS peptides, as previously reported [25].

### Statistical Analysis

Considering the absence of data and the exploratory aim of the trial, a convenient sample size of 30 patients was considered adequate to assess the safety of the study drug. Descriptive statistics were computed for all study variables. Categorical data were presented as numbers and percentages; continuous data were presented as mean (± standard deviation) or median [25th–75th percentiles], according to the distribution pattern of each variable, which was assessed using the Kolmogorov-Smirnov test prior to any calculations. To compare the means of the quantitative variables studied over time, the Generalized Estimation Equations (GEE Model) model was used. The main effect of the moment was tested by adjusting the results for the variable dose of the drug, since treated patients went over an increase in posology during the study period. The model was composed of an unstructured working correlation matrix, an estimator covariance matrix, and a normal distribution with identity link function. Analyzes were performed using IBM SPSS Statistics software, v.25. The significance level adopted was 0.05. Graphical analyzes were performed GraphPad Prism (version 9.3.1 for Macintosh, GraphPad Software, La Jolla, CA, US).

## Results

### Study population

Between August and December 2020, 29 patients were included; one patient was eventually excluded after acquiring previously unavailable information about active cancer disease, leaving 28 patients for the final analysis. The mean age of the study cohort was 55.8 ± 12.0 years and the mean body mass index (BMI) was 31.1 (± 7.3) Kg/m^2^; 13 patients (43.3%) were treated with chronic ACE inhibitors (ACEi) or Sartans (ARBs) treatment before admission. The baseline characteristics of the studied population are resumed in Table 1. The median ICU length of stay was 8.9 ± 8.7 days, and 6/28 (21.4%) patients died.

**Table 1:** Characteristics of the study population. Data are presented as mean (SD) or count (%)

| DEMOGRAPHICS |  |
| --- | --- |
| Age at inclusion, years | 55.9 ($\pm 12.5$) |
| Body Mass Index, Kg/m<sup>2</sup> | 31.1 ($\pm 7.3$) |
| Male gender, n | 22 (78.6) |
| IBGE classification, (n, %) |  |
| *White* | 10 (35.7) |
| *Black* | 2 (7.1) |
| *Brown* | 14 (50.0) |
| COMORBIDITIES |  |
| Arterial hypertension, n | 13 (46.4) |
| Chronic heart disease, n | 1 (3.6) |
| Diabetes, n | 10 (35.7) |
| Chronic neurological disorder, n | 1 (3.6) |
| Dementia, n | 1 (3.6) |
| Obesity*, n | 11 (39.2) |
| Chronic pulmonary disease, n | 1 (3.6) |
| Asthma, n | 2 (7.1) |
| Active smoking, n | 2 (7.1) |
| Chronic liver disease, n | 0 (0) |
| Other clinically relevant condition, n | 3 (10.7) |

| PARAMETERS AT INCLUSION |  |
| --- | --- |
| Temperature, °C | 36.7 (±0.8) |
| Heart rate, beats/min | 94.2 (±17.3) |
| Respiratory rate, /min | 23.8 (±5.6) |
| Systolic blood Pressure, mmHg | 137.2 (±19.1) |
| Diastolic Blood Pressure, mmHg | 79.0 (±12.7) |
| SaO<sub>2</sub>, % | 94.1 (±3.2) |
| FiO<sub>2</sub>, % | 52.0 (±27.5) |

IBGE= Instituto Brasileiro de Geografia e Statistica; SaO<sub>2</sub>= arterial oxygen saturation; FiO<sub>2</sub>= fraction of inspired oxygen. \*Defined as a body mass index >30.

### Ang-(1-7) administration and primary outcome

Median length of Ang-(1-7) infusion was 4.3 ± 2.3 days and dose escalation were possible in all patients but one. In two patients, the infusion was discontinued on multiple occasions because of technical problems, unrelated to SAEs. Eight (28.5%) patients received the treatment for 7 days.

No significant changes in the mean blood pressure or heart rate were recorded during the infusion, compared to the baseline values (Figure 1).

During the period of intervention, 5/28 (17.8%) patients required vasopressors; however, 4 of them had low dose norepinephrine (i.e. <0.05 mcg/kg*min, while one patient required higher doses 3 days after the initiation of Ang-(1-7) infusion and eventually died of septic shock. This event was considered unrelated to drug administration. Another SAE was reported during the study period; slightly after the increase in the infusion rate after 24 hours, one patient presented unexplained sinus bradycardia (i.e. heart rate < 50 beats per minute), without concomitant hypotension. This patient was under concomitant treatment with pilocarpine collyrium as chronic treatment for glaucoma. The attending physician decided to stop the infusion, as the SAE was possibly related to the study drug, and bradycardia resolved within few minutes and the infusion was uneventfully resumed at the rate of 5 mcg/Kg*day. This event was presented within 6 hours to the DSMC, which advised for not increasing the administration of Ang-(1-7) in this patient and considered bradycardia as potentially related to the study drug. No other expected SAEs were observed in these patients.

Despite no participants was on invasive mechanical ventilation at inclusion, nine patients (32.1 %) required this intervention during the study period, 5 (17.8%) requiring neuromuscular blocking agents and 6 (21.4%) required prone positioning. No patients required renal replacement therapy or ECMO during the infusion time.

### Secondary Outcomes

The OFDs and ICU free days were 19.1 ± 6.7 days and 17.5 ± 17.6 days, respectively. Total ventilator free days were 18.7± 13.2 days. Secondary infections were observed in 5 (17.8%) patients. One patient presented clinically relevant deep venous thrombosis. There was no significant increase over time of leukocytes, CRP and creatinine, despite an increase in blood urea concentration over the first 5 days, as presented in supplementary figure 1.

Two patients presented a significant increase in the plasmatic levels of Ang I and Ang II within 72 hours of Ang-(1-7) infusion. The time-based evolution of the RAS peptides is presented in Figure 2; no significant changes of circulating RAS peptides were observed over time, except a reduction of plasmatic levels of Ang-(1-7) between T0 and T72 and an increase of the Ang II / Ang I between T24 and T72.

**Figure 2:**
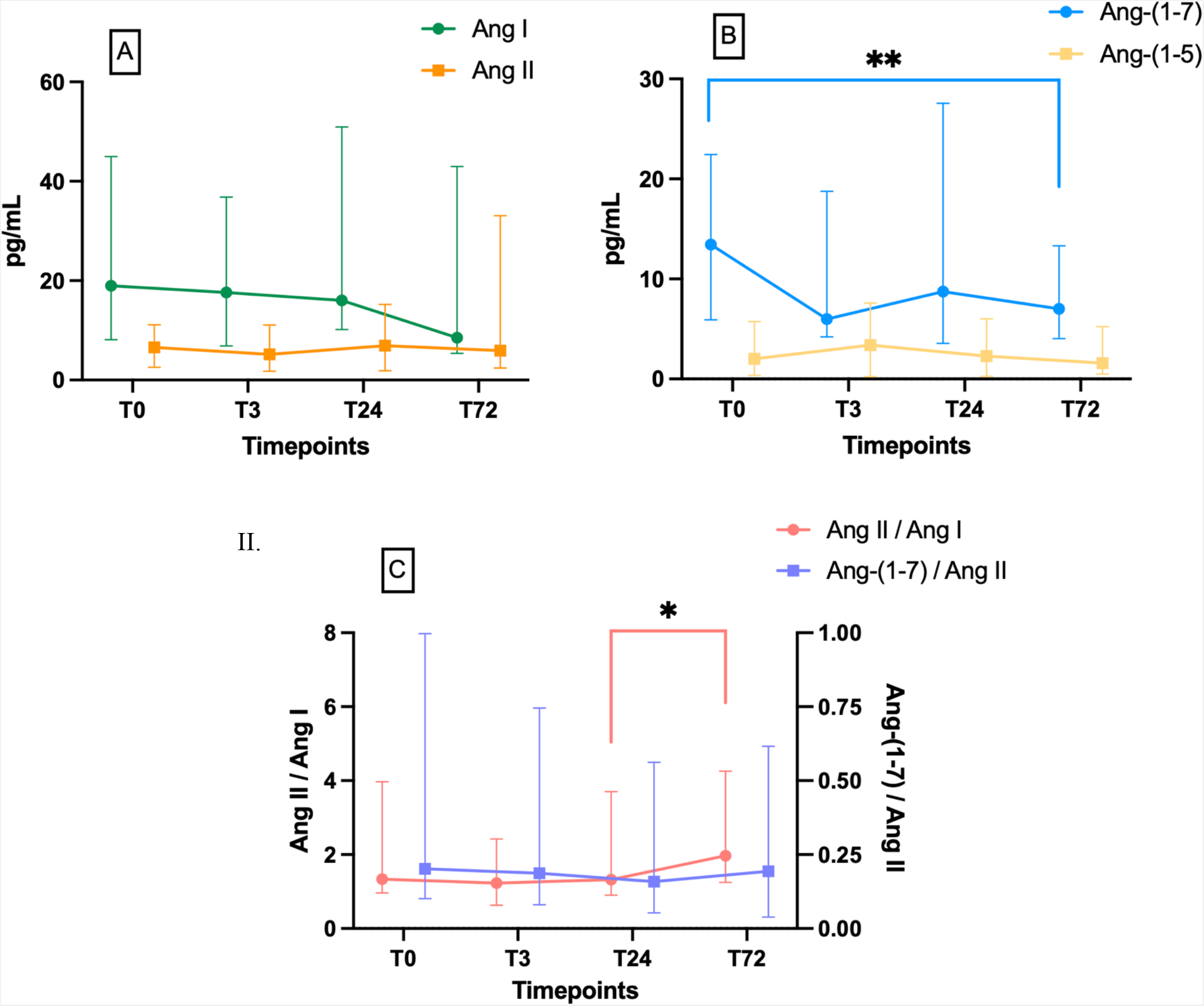
Time-course of RAS peptides. A- Angiotensin I and Angiotensin II. B-Angiotensin-(1-7). and Angiotensin-(1-5). C- Ang II/Ang I and Ang-(1-7)/Ang II. Symbols represents medians and bars represents interquartile range

## Discussion

In this Phase I clinical trial, we showed that intravenous infusion of Ang-(1-7) in severe COVID-19 patients requiring an ICU admission was safe. To our knowledge, this is the first human trial that has administered this compound intravenously for up 7 days and the first intravenous application of Ang-(1-7) in the context of COVID-19. Only one patient experienced a SAE (i.e., severe sinus bradycardia), which required infusion rate decrease, although no hemodynamic instability was recorded. The concomitant administration of the study drug with another medication providing cholinergic stimulation might have provided a synergistic effect and potentialized the bradycardic properties of pilocarpine.

In another study, Lissoni et al. tested the administration of Ang-(1-7) in COVID-19 patients [28]. In this Phase II study, Ang-(1-7) was administered orally in association with melatonin and cannabidiol to symptomatic non-hospitalized COVID-19 patients and their cohort was then matched with an historical control group. Importantly, no previous dose finding, nor safety assessment were conducted and the effects of Ang-(1-7) administration on the concentrations of RAS peptides were not measured. Moreover, the authors assumed that Ang-(1-7) was deeply downregulated in COVID-19 patients; however, two recent studies have shown higher Ang-(1-7) levels in COVID-19 patients, at least in those with more severe forms, when compared to healthy individuals [27, 28]. On the other hand, the fact that Ang-(1-7) was not suppressed at baseline does not exclude that a relative insufficiency could still be present in COVID-19 patients, especially at tissular level, and patients with a lower baseline Ang-(1-7) levels had longer ICU stay, higher mortality and have been found associated with disease severity. Interestingly, Ang-(1-7) tended to decrease in plasma during the infusion, with concomitant increase in plasmatic Ang-(1-5). Concomitantly, Ang II levels were within lower ranges at baseline and increased during drug administration. Those compelling results remain to be elucidated; however, we hypothesized that the exogenous Ang-(1-7) could have been rapidly metabolized during its passage through the pulmonary circulation, decreasing its concentration at the arterial sampling site with concomitant increase in Ang-(1-5). In humans, modulation of RAS during acute respiratory distress syndrome (ARDS) has already been investigated using a recombinant form of the ACE2 enzyme (rACE2), which resulted into decreased levels of Ang II and a parallel increase in Ang-(1-7), but without clear clinical benefit [29].

In this study, the administration of Ang-(1-7) was well tolerated. No significant hypotension was observed, as low dose norepinephrine was required in only 4 patients. Moreover, bradycardia was reported in one patient; a slight bradycardic effect of Ang-(1-7) has been observed in rats instrumented with continuous telemetry [30], and it well known that this peptide improves vagal tonus in hypertensive and normotensive rats [12]. We hypothesized that a potential interaction of the cholinergic drug pilocarpine with Ang-(1-7) might have explained bradycardia, although no demonstration of this phenomenon has been found. Clearly, these hemodynamic events, although well tolerated, might indicate the need for an accurate and continuous monitoring of such patients during Ang-(1-7) intravenous administration.

Our study has several limitations. First, as a Phase I clinical trial, particular attention needs to be used when interpreting clinical variables, because, due to the nature of the study, all patients received the study treatment. Moreover, only 29 out of the 30 initially planned inclusions were accomplished, due to technical problems. Nevertheless, these safety findings have provided a solid basis to conduct a phase II clinical study on this drug. Second, we did not measure all the components of the RAS, nor the enzymatic activities of ACE and ACE2 through the entire drug administration. Nevertheless, we have measured the plasmatic levels of several RAS peptides at multiple time-points using mass spectrometry, which provides solid and reproducible data. Third, we decided to include only patients with mild severity of the disease and with an age lower than 80 years old and thus we cannot directly extend our results to the general COVID-19 population admitted to the ICU. Fourth, we have tested only two dose regimens, and we cannot exclude that a higher dose might have resulted in more SAEs. Drug regimens were derived from experimental data, in which a clinically relevant effect was demonstrated. Lastly, we could not adjust our results on several confounders, such as the chronic use of ACEi or ARBs, which have relevant effects on RAS and might influence plasmatic peptides concentrations.

## Conclusions

Intravenous administration of Ang-(1-7) was safe in COVID-19 patients admitted to the ICU; the effectiveness of this drug will be tested in a Phase II clinical trials.

## Data Availability

The data that support the findings of this study are available from the corresponding author, [R.A.S.S.], upon reasonable request.

## List of Abbreviations

ACE: angiotensin converting enzyme
ACE2: angiotensin converting enzyme 2
ACEi: ACE inhibitors
ARBs: angiotensin II receptor blockers, or sartans
Ang I: angiotensin I
Ang-(1-7): angiotensin-(1-7)
Ang II: angiotensin II
ICU: intensive care unit
OFDs: oxygen-free days
RAS: renin angiotensin system
SAEs: serious adverse events
DSMC: data safety monitoring committee

## Declarations

### Ethics approval and consent to participate

The study was approved by the ethic committee of the Federal University of Minas Gerais (CAAE: 34080720.0.1001.5149) and was carried out in accordance with the declaration of Helsinki after the approval of the CONEP (national council of ethics in research).

Written informed consent was obtained from a legal representative or the patient, depending on the circumstances, before inclusion.

### Funding

The study was conducted with the support of the Université Libre de Bruxelles (ULB) grant for COVID-19 (Research Number 5C06G000010), Fundação de Amparo a Pesquisa do Estado de Minas Gerais (Fapemig), Angitec (Brazilian Start-up). Dr Annoni received research grant from Fonds Erasme pour la Recherche Médicale.

### Authors contributions

F.A., A.V.M., R.A.S.S. and F.S.T. have conceived the study and draft the initial protocol. A.V.M., F.A.S., L.B.R, G.C.O., A.H.D.S.M, M.C.J.H., B.D.P., N.A.R., D.A.A.P, M.M.L.D., T.G.F.S., A.C.C, J.C.M., D.M.S., M.J.C.S. included all patients and entered the data in the database. T.V.B., F.A. and R.A.S.S. performed the statistical analysis. F.A., A.V.M, R.A.S.S. and F.S.T were responsible for a first draft of the manuscript. F.A. prepared all figures and tables. All authors reviewed the manuscript.

## Conflict of Interests

The authors of the trial declared to have no conflict of interest. Prof Robson is the CEO of the company Angitec, which provided economic and logistic support without any monetary or contractual compensation.

## Notes

### Clinical Trial

Registro Brasileiro de Ensaios Clinicos, UTN code: U1111-1255-7167, registered on 08/05/2020; ClinicalTrials.gov Identifier: NCT04633772

### Funding Statement

The study was conducted with the support of the Universite Libre de Bruxelles (ULB) grant for COVID-19 (Research Number 5C06G000010), Fundacao de Amparo a Pesquisa do Estado de Minas Gerais (Fapemig), Angitec (Brazilian Start-up). Dr Annoni received research grant from Fonds Erasme pour la Recherche Medicale.

### Author Declarations

The study was approved by the ethic committee of the Federal University of Minas Gerais (CAAE: 34080720.0.1001.5149) and was carried out in accordance with the declaration of Helsinki after the approval of the CONEP (national council of ethics in research). Written informed consent was obtained from a legal representative or the patient, depending on the circumstances, before inclusion.

